# Epidemiological Trends of Coronavirus Disease 2019 in China

**DOI:** 10.1101/2020.03.13.20035642

**Authors:** Bilin Chen, Huanhuan Zhong, Yueyan Ni, Lulu Liu, Jinjin Zhong, Xin Su

## Abstract

**Background:** The Coronavirus Disease 2019 (COVID-19) epidemic broke out in Wuhan, China, and it spread rapidly. Since January 23, 2020, China has launched a series of unusual and strict measures, including the lockdown of Wuhan city to contain this highly contagious disease. We collected the epidemiological data to analyze the trend of this epidemic in China.

**Methods:** We closely tracked the Chinese and global official websites to collect the epidemiological information about COVID-19. The number of total and daily new confirmed cases of COVID-19 in China was presented to illustrate the trend of this epidemic.

**Results:** On January 23, 2020, 835 confirmed COVID-19 cases were reported in China. On February 6, 2020, there were 31211 cases. By February 20, 2020, the number reached as high as 75,993. Most cases were distributed in and around Wuhan, Hubei province. Since January 23, 2020, the number of daily new cases in China except Hubei province reached a peak of 890 on the eleventh day and then it declined to a low level of 34 within two full-length incubation periods (28 days), and the number of daily new cases in Hubei also started to decrease on the twelfth day, from 3156 on February 4, 2020 to 955 on February 15, 2020.

**Conclusion:** The COVID-19 epidemic has been primarily contained in China. The battle against this epidemic in China has provided valuable experiences for the rest of the world. Strict measures need to be taken as earlier as possible to prevent its spread.

## INTRODUCTION

Emerging infectious diseases are great threats to public health worldwide. Towards the end of the year 2019, an outbreak of a novel coronavirus (Severe Acute Respiratory Syndrome Coronavirus 2, SARS-CoV-2)(1-3) infection occurred in Wuhan, Hubei Province, China. It rapidly spread to every corner of China and many other places around the world. SARS-CoV-2 is a new member of the coronavirus family, which is different to Severe Acute Respiratory Syndrome Coronavirus, SARS-CoV.(4) Current reports show that SARS-CoV-2 is more contagious than SARS-CoV. The new coronavirus pneumonia has been named as Coronavirus Disease 2019 (COVID-19) by the World Health Organization.(5)

On February 20, 2020, the total number of confirmed COVID-19 cases had reached as high as 75,993 in China. Among these cases, 2239 died, and most of the deaths occurred in elderly patients with certain underlying illnesses.(6-8) To confront this terrible epidemic, China took restrictive measures to prevent the spread of SARS-CoV-2. Restricting the population movement, shutting down of schools and factories, building up new shelter hospitals, and many other measures were carried out. In addition to ordinary measures, the lockdown of Wuhan since January 23, 2020, a city with more than 10 million people, was a unique and unusual method. At the same time, many provinces and cities in China initiated a level I public health event response. To further control the epidemic, from February 5th, 2020, all confirmed and suspected COVID-19 patients and close contacts in Wuhan were asked to quarantine at assembly sites instead of home quarantine. What is the effect of these counter-measures for controlling this epidemic?

The National Health Commission of the People’s Republic of China (NHC) and The Center for Disease Control and Prevention of China (CDC) have publicly shared detailed epidemiological information about COVID-19 every day since January 20, 2020.(7, 8) Based on the information and published studies about COVID-19, many scholars have expressed different opinions on the effect of these measures and the trend of this epidemic. Michael Levitt, a professor of Stanford University, speculated that since January 25, 2020, the national death toll and the death toll in Hubei province showed a monotonous declining trend. The external linear correlation suggests that the number of new deaths in the coming week will decline rapidly, leading to the conclusion that the epidemic would terminate soon.(9)

However, there are many other different opinions. The University of Lancaster, the University of Florida, and the Centre for Viral Research at the University of Glasgow carried out modeling analysis based on the data before January 21, 2020. They estimated that the number of infected individuals may be as high as 14464 cases on January 22, 2020 in Wuhan. If there were no effective measures, the total number of infection cases would reach as high as 105077 on January 29, 2020.(10) A team of researchers from Xi’an Jiaotong University in China and York University in Canada worked together to build a more realistic Susceptible-Exposed-Infectious-Recovered (SEIR) model, and the novel coronavirus infection could reach an inflection point on March 10, 2020.(11)

Big gaps existed between different predictions. Which one is identical with the actual situation? In addition, being the eye of the storm, Wuhan city and Hubei Province show a different situation compared to that in the other provinces in China. Therefore, we compared the data of different places to predict the trend of this epidemic in China.

## METHODS

### Sources of data and searches

According to the study of early epidemiology of COVID-19 transmission, the incubation period of COVID-19 extends from 1 to 14 days.(12) Therefore, we collected data of 28 days (two full-length incubation periods) to analyze the trend of COVID-19 in China. We closely tracked the relevant resources including Chinese and global official websites and announcements between January 23, 2020 and February 20, 2020. China’s data are based on the everyday briefing by the NHC(7) and CDC(8). The relevant data included the provincial distribution of the epidemic on each day from January 23, 2020 to February 20, 2020.

### Case Definitions

According to the diagnostic and treatment protocol for COVID-19 (Trial version 5 revision) released by the NHC(13), confirmed cases were patients with laboratory-confirmed SARS-CoV-2 by real-time RT-PCR or next-generation sequencing. Comprehensive analysis of the epidemiological history, clinical manifestations, and evidence of virus nucleic acid was required. Clinically diagnosed cases were patients with a definite epidemiological history and typical clinical manifestations.

### Statistical analysis

Retrieved data were recorded in Microsoft Excel for Windows (version 18.19) for analysis. Continuous variables included the total number of confirmed cases daily, the number of newly confirmed cases, the number of deaths, and the number of severe and critical cases. Graph pad version 8.0.1 (GraphPad Software, America), Tableau version 2019.4.3 (Tableau Software, America), and Xmind version 2.0.2 (Xmind Ltd, China) were used for cartography. Data from 23 January 2020 to 20 February 2020 were utilized for China map drawing.

## RESULTS

COVID-19 was distributed in every province of China. Most of the cases were distributed in and around Wuhan city, China. As shown in Figure 1, the number of COVID-19 cases increased rapidly. From January 23, 2020 to February 20, 2020, the total number of cases increased from 835 to 75,993 in just 28 days. Most cases were found in the worst-hit areas of Hubei province. In Hubei, more cases were identified between February 7, 2020 and February 20, 2020. The epidemic spread rapidly from Hubei province to adjacent provinces and the whole country. In other provinces except Hubei, most cases were identified between January 24, 2020 and February 6, 2020.

**Figure 1.**
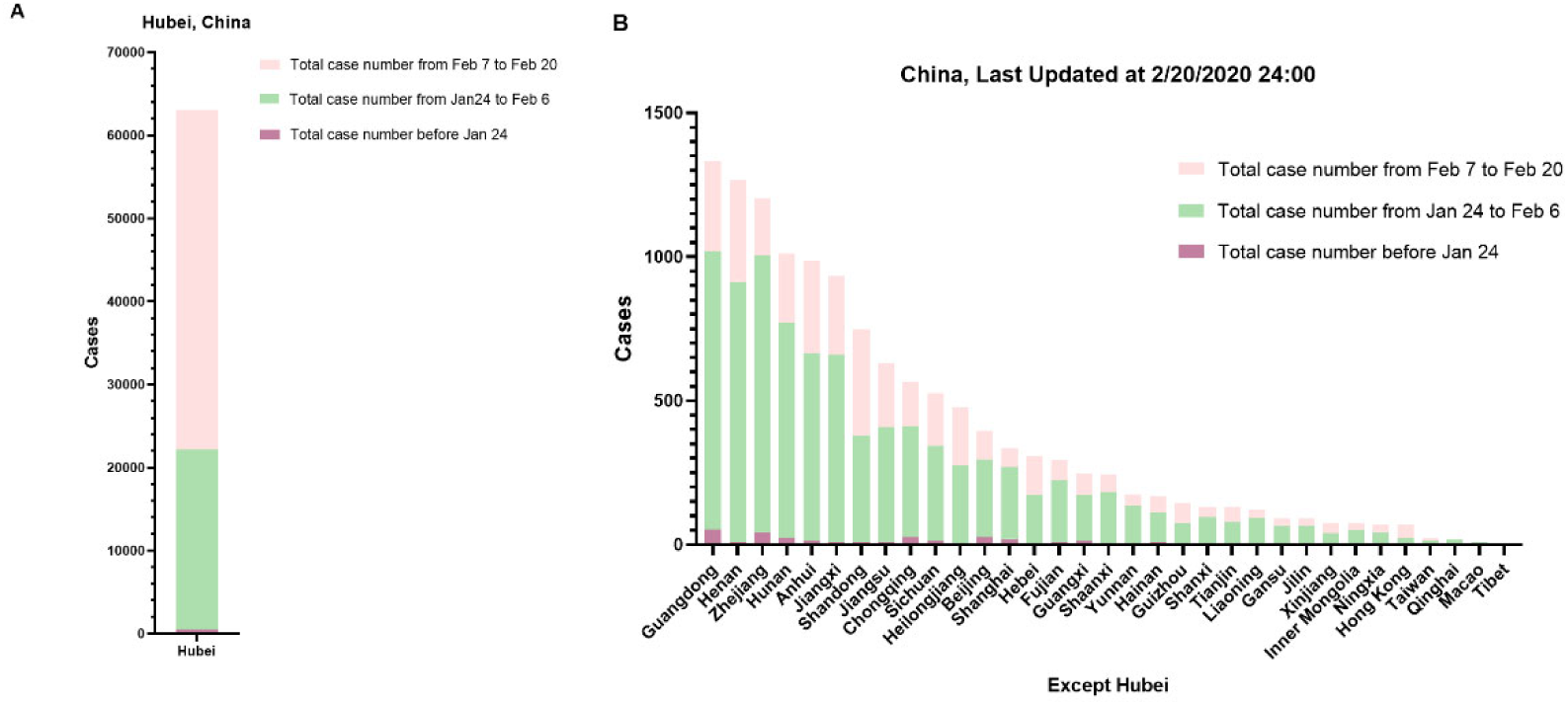
The number of confirmed cases in each province or region of China. Data before January 24, 2020 is presented in purple, data from January 24, 2020 to February 6, 2020 is presented in green, and data from February 7, 2020 to February 20, 2020 is presented in pink. 1A, the data of cases in Hubei Province, including clinically diagnosed cases after February 13, 2020. 1B, the data of confirmed cases in China except Hubei.

Since January 23, 2020, Wuhan, the capital city of Hubei Province, was in lockdown. Many other provinces responded to the public health incidents by adopting quarantine measures and community-level monitoring to control the epidemic spread. Figure 2 shows the changes in the cumulative number of confirmed cases and the daily number of new confirmed cases since the lockdown of Wuhan, Hubei province, on January 23, 2020.

**Figure 2.**
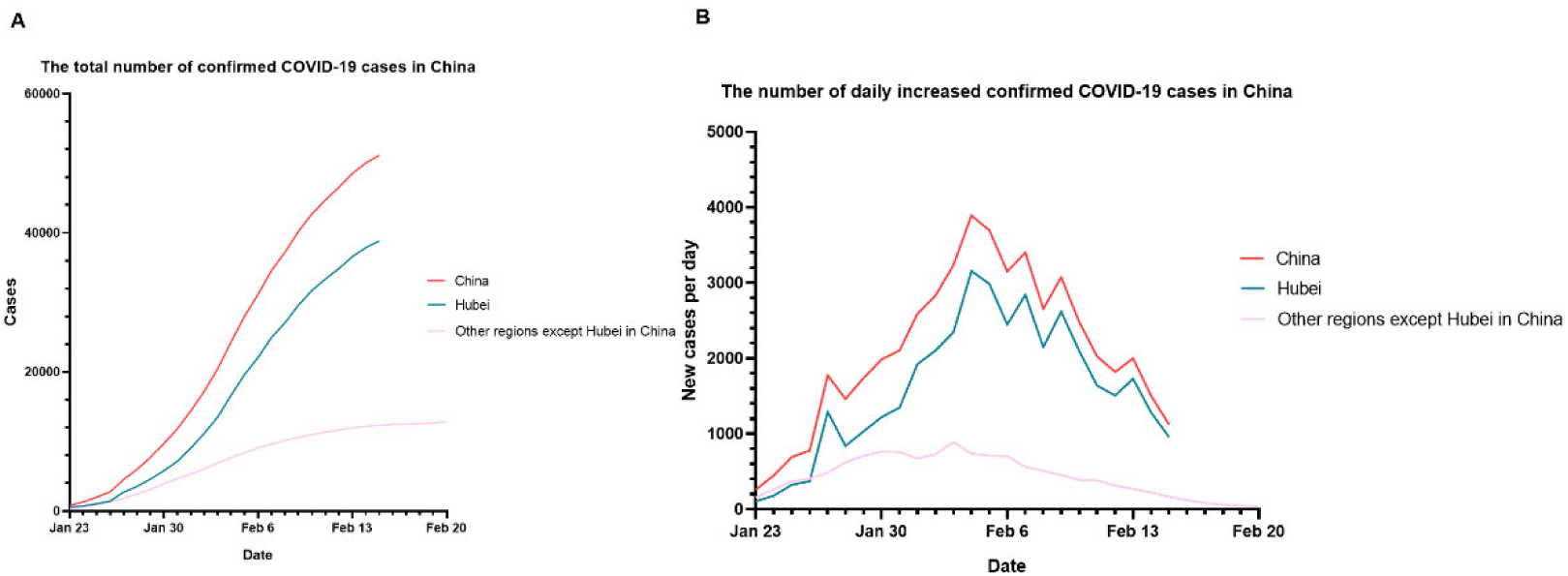
The data of the number of confirmed cases in China between January 23, 2020 at 24 pm and February 20, 2020 at 24 pm (Beijing time), (2A). The total number of confirmed COVID-19 cases in China, (2B). The number of daily increased confirmed COVID-19 cases in China. From February 16, 2020, in Hubei Province of China, the number of confirmed cases and clinical diagnosed cases were calculated together and reported as one number. Thus, the total number of cases for Hubei and China were presented till February 15, 2020.

Figure 2A shows that the cumulative number of confirmed cases kept increasing. Most of the cases were found in Hubei Province. Total number of confirmed cases outside Hubei province in China was 12,905 on February 20, 2020. Total number of confirmed cases in Hubei province was 38,839 on February 15, 2020. Figure 2B shows the daily change in the number of new confirmed cases since January 23, 2020. The daily number of new confirmed cases in most provinces of China showed a downward trend, including Hubei Province. The number of increased cases in the other provinces in China was significantly lower than that in Hubei province. The number of daily increased cases in the other provinces in China reached a peak of 890 on February 3, 2020, and then it continued to decline to as low as 34 on February 20, 2020. The number of daily increased cases in Hubei province began to decline on February 4, 2020.

The number of daily new cases in the other provinces except Hubei in China is presented in Figure 3. The number of daily new confirmed cases in 25 provinces shared almost the same obvious trend, increasing before February 5, 2020 and then declining. These provincial units are listed in the order of the total case numbers as follows: Guangdong, Henan, Zhejiang, Hunan, Anhui, Jiangxi, Shandong, Jiangsu, Chongqing, Sichuan, Heilongjiang, Beijing, Shanghai, Hebei, Fujian, Guangxi, Shaanxi, Yunnan, Hainan, Guizhou, Shanxi, Tianjin, Liaoning, Gansu, and Jilin.

**Figure 3.**
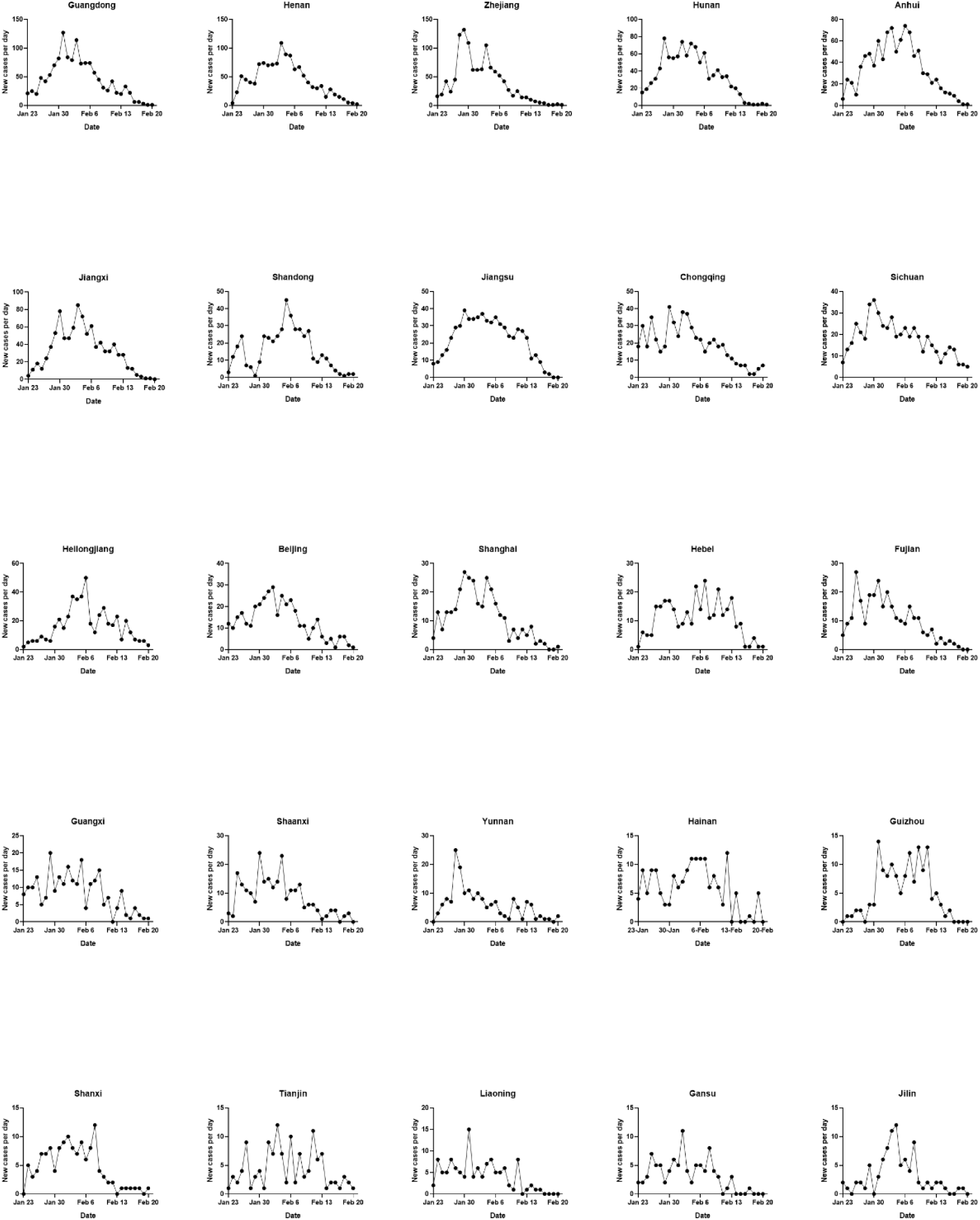
The trends of daily new confirmed cases in the top 25 provincial units in terms of the total cases over 90 were presented (The trend of Hubei has been presented in Figure 2B), including Guangdong, Henan, Zhejiang, Hunan, Anhui, Jiangxi, Shandong, Jiangsu, Chongqing, Sichuan, Heilongjiang, Beijing, Shanghai, Hebei, Fujian, Guangxi, Shaanxi, Yunnan, Hainan, Guizhou, Shanxi, Tianjin, Liaoning, Gansu, and Jilin. The trends in each province were presented based on the data between January 23, 2020 at 24 pm and February 20, 2020 at 24 pm (Beijing time).

After Hubei province, the four provinces with the highest number of COVID-19 cases in China were Guangdong, Henan, Zhejiang, and Hunan province. Figure 4 shows the time curves of the total confirmed cases and daily new cases in these four provinces. The overall trend of the total number of confirmed cases and the number of daily new cases in the four provinces showed almost the same trend. About 14 days (one incubation period) later, the number of daily new cases began to decline. After 28 days (two incubation periods), the number of daily new confirmed cases dropped to single digits.

**Figure 4.**
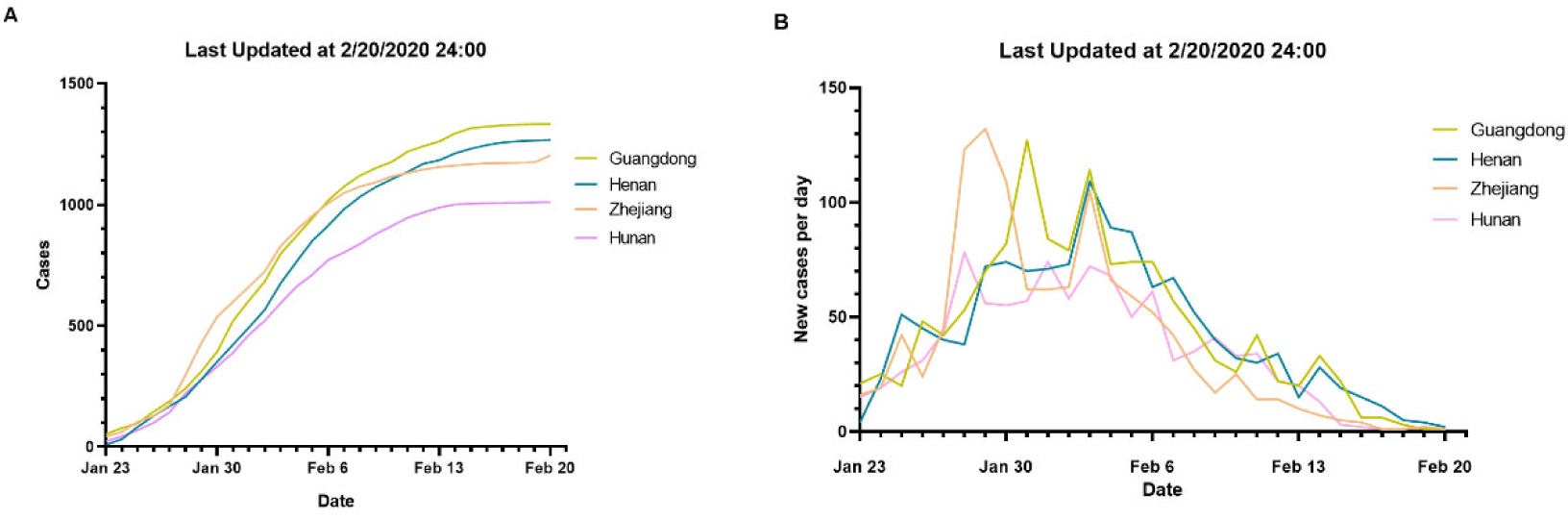
Changes in confirmed cases in four provinces of Guangdong, Henan, Zhejiang, and Hunan province; data between January 23, 2020 at 24 pm and February 20, 2020 at 24 pm (Beijing time), (4A) The total number of confirmed COVID-19 cases in Zhejiang, Guangdong, Henan, and Hunan province. (4B) The number of daily increased confirmed COVID-19 cases in Guangdong, Henan, Zhejiang, and Hunan province.

## DISCUSSION

The COVID-19 emergency response epidemiology team of CDC in China conducted an epidemiological analysis on COVID-19 and pointed out that since the first case was admitted to hospital in Wuhan in December, the outbreak had developed rapidly.(14) Based on the current reports, although the mortality rate is lower than that of SARS and Middle East Respiratory Syndrome (MERS), COVID-19 appears to be more infectious.(15-17) The above studies suggest that stricter measures need to be taken to contain the COVID-19 epidemic.

Realizing this critical situation, China adopted an old fashioned approach but the response in China changed over time to science and risk based approach, and China adopted a bold approach. The specific event timeline is shown in Figure 5. The data in this paper were released after the implementation of strict measures on January 23, 2020. The trends in these figures suggested that the epidemic had been contained in the two incubation periods after the emergency response was upgraded to the highest level and strictly implemented around China.

**Figure 5.**
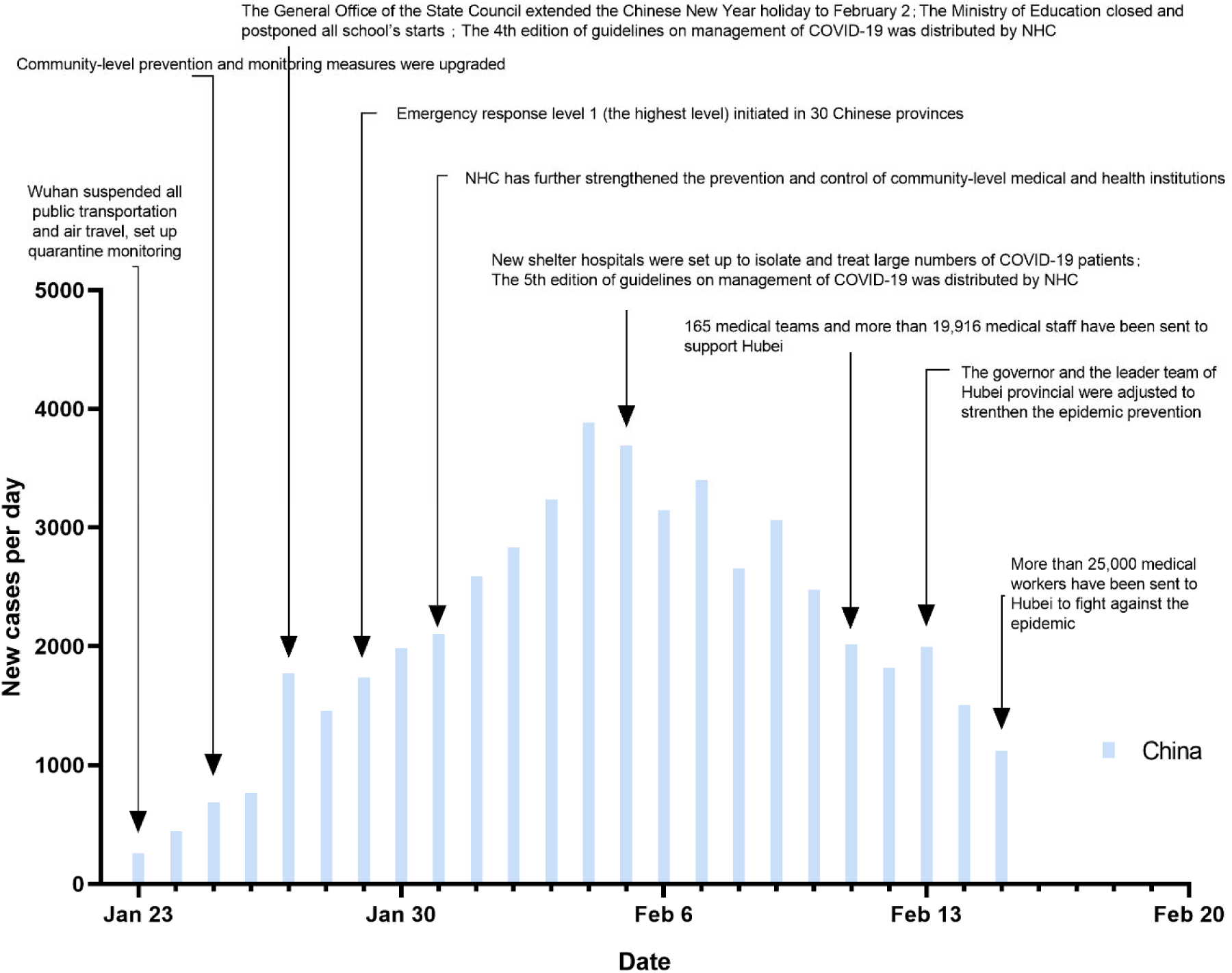
Major prevention and control measures in China since January 23, 2020.

The major prevention and control measures are shown in Figure 5; as the outbreak spread and the death toll increased, the Chinese government and healthcare authorities implemented unprecedented measures. Since January 23, 2020, Wuhan was in lockdown. All public transportation was suspended, The whole city was quarantined and monitored.(18) Soon after, these measures were extended to the remaining of Hubei Province and many other provinces in China.

Since the lockdown of Wuhan city, the shift of population from Wuhan city and Hubei province to the other parts of China ceased. The Chinese New Year holiday was extended.(19) The Ministry of Education postponed the school opening.(20) Meanwhile, the Ministry of Civil Affairs and the NHC further carried out community-level prevention in urban and rural areas.(21-23)

Figure 5 also shows that since January 23, 2020, the peak of new confirmed cases daily occurred from February 4 to February 7. The report of the WHO-China Joint Mission on COVID-19 indicates that according to the date of onset, the confirmed cases peaked between January 23 and January 27.(24) The peak of confirmed cases occurred 11 to 12 days later than the peak of onset, which is almost the length of the incubation period.

Although the total number of cases in Hubei province is still rising, the trend has begun to slow down, and the number of daily new cases in China has decreased significantly. Other than Hubei, four provinces of Zhejiang, Guangdong, Henan, and Hunan had the highest number of COVID-19 cases (Figure 4). In one infectious incubation period (14 days), the number of daily new cases reached the peak, and it decreased continuously in the second incubation period (14–28 days).

After the adoption of strict prevention and control measures on January 23, 2020, the daily number of new confirmed cases decreased significantly in the two infectious incubation periods (28 days). The epidemic reached its peak earlier in most provinces than that suggested by many prediction models. This shows that China’s aggressive approach has changed the course of the epidemic.

On comparing Figure 2A with 2B, we found that the number of newly confirmed cases in Hubei province decreased significantly since February 6, 2020. The increase in confirmed cases also slowed down. Since the opening of a shelter hospital in Hubei province around February 5th and strengthening of the community-level quarantine, the transmission from person to person was further inhibited. After two incubation periods, the number of existing confirmed cases began to decline.

In summary, COVID-19 is quite different from SARS. It is even more infectious and destructive. There are still many uncertainties about the epidemic. After the adoption of aggressive measures, China has contained the epidemic. However, the cost is huge. Millions of healthcare workers and social workers have devoted themselves in the fight against this epidemic. Some of them have even dedicated their lives. Currently, the number of COVID-19 cases is increasing in many other countries. The battle against COVID-19 in China has provided many valuable experiences for the rest of the world.

## Data Availability

Publicly available datasets were analyzed in this study. This data can be found here:
[http://www.nhc.gov.cn/xcs/yqtb/list_gzbd.shtml and
https://www.who.int/emergencies/diseases/novel-coronavirus-2019/situation-reports].

https://www.who.int/emergencies/diseases/novel-coronavirus-2019/situation-reports

## AUTHOR CONTRIBUTORS

Xin Su had the conception for and designed the study, and take responsibility for the integrity of the data and the accuracy of the data analysis. Bilin Chen, Huanhuan Zhong, Yueyan Ni, Lulu Liu and Jinjin Zhong contributed to the data acquisition. Xin Su and Bilin Chen contributed to the statistical analysis and interpretation. All authors contributed to data acquisition, data analysis, or data interpretation, and reviewed and approved the final version.

## DATA AVAILABILITY STATEMENT

Publicly available datasets were analyzed in this study. This data can be found here: [http://www.nhc.gov.cn/xcs/yqtb/list_gzbd.shtml and https://www.who.int/emergencies/diseases/novel-coronavirus-2019/situation-reports].

## ETHICS STATEMENT

Ethical approval for this study and written informed consent from the participants of the study were not required in accordance with local legislation and national guidelines.

## ACKNOWLEDGMENTS

This work was supported by the Project of Natural Science Foundation of China [Grant number 81873400], the Key Project of Jiangsu Commission of Health (K2019004), and “333 project” of Jiangsu Province (BRA2019339). We thank LetPub (www.letpub.com) for its linguistic assistance during the preparation of this manuscript.

## REFERENCE

1. Wuhan Municipal Health Commission. Report of clusterring pneumonia of unknown etiology in Wuhan City. Wuhan, China: Wuhan Municipal Health Commission (2019) [cited 2019 December 31]. Available from: http://wjw.wuhan.gov.cn/front/web/showDetail/2019123108989.

2. Pneumonia of unknown cause – China Geneva, Switzerland: World Health Organization (2020) [cited 2020 January 12]. Available from: https://www.who.int/csr/don/05-january-2020-pneumonia-of-unkown-cause-china/en/.

3. Gorbalenya AE, Baker SC, Baric RS, de Groot RJ, Drosten C, Gulyaeva AA, et al. Severe acute respiratory syndrome-related coronavirus: The species and its viruses – a statement of the Coronavirus Study Group. *bioRxiv* (2020):2020.02.07.937862. doi: 10.1101/2020.02.07.937862.

4. Lu R, Zhao X, Li J, Niu P, Yang B, Wu H, et al. Genomic characterisation and epidemiology of 2019 novel coronavirus: implications for virus origins and receptor binding. The Lancet (2020). doi: 10.1016/S0140-6736(20)30251-8.

5. WHO Director-General’s remarks at the media briefing on 2019-nCoV on 11 February 2020 Geneva, Switzerland: World Health Organization (2020) [cited 2020 February 11]. Available from: https://www.who.int/dg/speeches/detail/who-director-general-s-remarks-at-the-media-briefing-on-2019-ncov-on-11-february-2020.

6. Coronavirus disease (COVID-2019) situation reports Geneva, Switzerland: World Health Organization (2020) [cited 2020 February 22]. Available from: https://www.who.int/emergencies/diseases/novel-coronavirus-2019/situation-reports.

7. National Health Commission’s briefing on the pneumonia epidemic situation. (2020) [cited 2020 February 12]. Available from: http://www.nhc.gov.cn/xcs/yqtb/list_gzbd.shtml.

8. Chinese Center for Disease Control and Prevention’s briefing on the pneumonia epidemic situation.: Chinese Center for Disease Control and Prevention (2020) [cited 2020 February 20]. Available from: http://www.chinacdc.cn/jkzt/crb/zl/szkb_11803/jszl_11809/.

9. Nobel laureate predicts Wuhan outbreak may be close to end: Sina science and technology (2020) [cited 2020 February 3]. Available from: https://mbd.baidu.com/newspage/data/landingshare?pageType=1&isBdboxFrom=1&context=%7B%22nid%22:%22news_9947297270045930268%22,%22sourceFrom%22:%22bjh%22%7D&_refluxos=i3&from=singlemessage&isappinstalled=0.

10. Read JM, Bridgen JRE, Cummings DAT, Ho A, Jewell CP. Novel coronavirus 2019-nCoV: early estimation of epidemiological parameters and epidemic predictions. *medRxiv* (2020):2020.01.23.20018549. doi: 10.1101/2020.01.23.20018549.

11. Tang B, Wang X, Li Q, Bragazzi NL, Tang S, Xiao Y, et al. Estimation of the Transmission Risk of the 2019-nCoV and Its Implication for Public Health Interventions. J Clin Med (2020) 9(2):E462. doi: 10.3390/jcm9020462. PubMed PMID: 32046137.

12. Li Q, Guan X, Wu P, Wang X, Zhou L, Tong Y, et al. Early Transmission Dynamics in Wuhan, China, of Novel Coronavirus-Infected Pneumonia. N Engl J Med (2020):10.1056/NEJMoa2001316. doi: 10.1056/NEJMoa2001316. PubMed PMID: 31995857.

13. Diagnostic and treatment protocol for Novel Coronavirus Pneumonia (Trial version 5 revision): General Office of National Health Commission, General Office of National Administration of Traditional Chinese Medicine (2020). Available from: http://www.nhc.gov.cn/xcs/zhengcwj/202002/d4b895337e19445f8d728fcaf1e3e13a/files/ab6bec7f93e64e7f998d802991203cd6.pdf.

14. Novel Coronavirus Pneumonia Emergency Response Epidemiology T. The epidemiological characteristics of an outbreak of 2019 novel coronavirus diseases (COVID-19) in China. Zhonghua Liu Xing Bing Xue Za Zhi (2020) 41(2):145–51. doi: 10.3760/cmaj.issn.0254-6450.2020.02.003. PubMed PMID: 32064853.

15. Huang C, Wang Y, Li X, Ren L, Zhao J, Hu Y, et al. Clinical features of patients infected with 2019 novel coronavirus in Wuhan, China. The Lancet (2020) 395(10223):497–506. doi: 10.1016/S0140-6736(20)30183-5.

16. Paules CI, Marston HD, Fauci AS. Coronavirus Infections-More Than Just the Common Cold. JAMA (2020):10.1001/jama.2020.0757. doi: 10.1001/jama.2020.0757. PubMed PMID: 31971553.

17. Wang C, Horby PW, Hayden FG, Gao GF. A novel coronavirus outbreak of global health concern. The Lancet (2020) 395(10223):470–3. doi: https://doi.org/10.1016/S0140-6736(20)30185-9.

18. Announcement from the Headquarter for novel coronavirus pneumonia prevention and control (No 1). Beijing: China National Health Commission (2020) [cited 2020 January 24]. Available from: http://www.gov.cn/xinwen/2020-01/23/content5471751.htm.

19. Notice of the General Office of the State Council on the extension of the Spring Festival holiday in 2020: The General Office of the State Council (2020) [cited 2020]. Available from: http://www.gov.cn/zhengce/content/2020-01/27/content5472352.htm.

20. Notice of the Ministry of Education on the postponement of the start of the spring semester in 2020: The Ministry of Education (2020) [cited 2020 January 27]. Available from: http://www.gov.cn/zhengce/zhengceku/2020-01/28/content5472571.htm.

21. Novel coronavirus pneumonia prevention and control program (version 3): General Office of National Health Commission (2020) [cited 2020 January 28]. Available from: http://www.nhc.gov.cn/xcs/zhengcwj/202001/470b128513fe46f086d79667db9f76a5/files/8faa1b85841f42e8a0febbea3d8b9cb2.pdf.

22. Novel coronavirus pneumonia prevention and control program (version 4): General Office of National Health Commission (2020) [cited 2020 February 7]. Available from: http://www.nhc.gov.cn/xcs/zhengcwj/202002/573340613ab243b3a7f61df260551dd4/files/c791e5a7ea5149f680fdcb34dac0f54e.pdf.

23. Novel coronavirus pneumonia prevention and control program (version 5): General Office of National Health Commission (2020) [cited 2020 February 21]. Available from: http://www.nhc.gov.cn/xcs/zhengcwj/202002/a5d6f7b8c48c451c87dba14889b30147/files/3514cb996ae24e2faf65953b4ecd0df4.pdf.

24. Report of the WHO-China Joint Mission on Coronavirus Disease 2019 (COVID-19): World Health Organization (2020) [cited 2020 February 29]. Available from: https://www.who.int/docs/default-source/coronaviruse/who-china-joint-mission-on-covid-19-final-report.pdf.

